# PED-X-Bench: A Benchmark of Adult-to-Pediatric Extrapolation Decisions in FDA Drug Labels

**DOI:** 10.1101/2025.05.22.25328187

**Authors:** Apoorva Srinivasan, Jacob Berkowitz, Nadine A. Friedrich, Kevin Tsang, Aditi Kuchi, José Acitores, Michael Zietz, Ryan S. Czarny, Hongyu Liu, Nicholas P. Tatonetti

## Abstract

Pediatric trials are ethically and logistically difficult, so the U.S. FDA often extrapolates adult data to children when justified. Yet no public resource systematically documents these decisions. We present **PED-X-Bench**, the first dataset and benchmark that encodes FDA pediatric-extrapolation outcomes as a four-way classification task (*Full, Partial, None, Unlabeled*). PED-X-Bench contains 737 FDA drug-label sections (≈ 1 M words of source text) for approvals issued 2007–2024 across all therapeutic areas. A two-stage *o3-mini* prompting pipeline mined full FDA label text; nine domain reviewers then adjudicated a stratified sample of 135 labels yielding an accuracy F1 of 0.74 and 0.63 respectively (inter-annotator κ = 0.678) and spot-checking the remainder. For every drug we release the ground-truth label, concise efficacy and pharmacokinetic/safety summaries, and harmonized study metadata. To showcase utility we release two baseline models: (i) a logistic-regression classifier that uses structured metadata from FDA’s pediatric-labeling dataset, and (ii) a fine-tuned BigBird BERT that ingests full label text. Both baselines perform modestly, leaving ample headroom for future work. PED-X-Bench enables research on pediatric drug development, clinical NLP and drug safety; dataset card and code are made available here: github.com/tatonetti-lab/PedXBench huggingface.co/datasets/apoorvasrinivasan/Ped-X-Bench

## 1 Introduction

Despite tremendous efforts over the past decades, off-label medicine use in pediatric populations are still at an undesirable level of 40 %, overall, and up to 90 % in neonates [1]. Because growth and maturation reshape both pharmacokinetics and disease biology, transplanting adult evidence into children can yield sub-therapeutic dosing, diminished efficacy, or an elevated risk of adverse reactions [2–4]. The underlying cause is straightforward yet hard to remedy: well-controlled pediatric trials face small sample sizes, heightened ethical scrutiny, and enormous developmental heterogeneity, all of which inflate cost and complexity and hamper recruitment.

Recognizing these issues, two pieces of legislation were passed in the US, the Best Pharmaceuticals for Children Act (BPCA) and the Pediatric Research Equity Act (PREA) [5, 6]. These laws seek to reduce off-label pediatric prescribing by requiring and/or providing financial incentives for drug manufacturers to perform pediatric clinical trials. BPCA is often referred to as the “carrot” because it offers manufacturers six months of additional market exclusivity in exchange for conducting pediatric trials. Whereas PREA is often referred to as the “stick” because it requires manufacturers in many cases to conduct pediatric trials in order to receive FDA approval for adult use. These initiatives have led to over 800 pediatric labeling changes [7], dramatically increasing the number of drugs with age-specific safety, efficacy, and dosing guidance for children. Yet, many approved drugs still rely heavily on extrapolation: the use of adult efficacy, safety, and pharmacokinetic (PK) data to support pediatric indications.

In 2024, the FDA adopted the ICH E11A guideline [8], which provides a harmonized, step-wise framework for pediatric extrapolation. This guidance emphasizes developing a structured “extrapolation concept” based on disease similarity, drug pharmacology, and response to treatment between reference and target populations. Importantly, it re-frames extrapolation not as a discrete yes-or-no decision, but as a continuum—recognizing that the degree of similarity, the level of uncertainty, and the required evidence can vary substantially across indications and populations.

Yet from a computational perspective, this continuum presents a challenge: it lacks standardization, clear boundaries, and supervision signals for training models. While clinical decisions may operate in gray areas, machine learning systems, especially those trained on textual regulatory data, benefit from structured, consistent labels to learn predictive patterns and support reproducibility. Moreover, historical FDA regulatory decisions have, in practice, been interpreted and applied in categorical terms: sponsors often receive labeling feedback that effectively falls into coarse bins such as *full, partial*, or *none* in terms of extrapolation acceptability. [9]. The categorized approach, e.g., full, partial, and no extrapolation, was based on a flowchart evaluation published in FDA draft guidance 2014. [10]

Despite the centrality of extrapolation to pediatric drug development, to date, no public dataset has systematically annotated or categorized extrapolation decisions in FDA drug labels. Previous work has involved manual review of extrapolation of a relatively small number of pediatric studies from internal FDA documents which are not publicly available [11]. On the other hand, recent NLP work on FDA labels targets toxicity or adverse-event extraction [12, 13] or general summarisation [14], not extrapolation. Thus, neither a formal, historically grounded schema nor an automated method for inferring these decisions from text exists. We present **PED-X-Bench** to fill this gap: it couples a four-way extrapolation taxonomy with LLM-generated rationales across 737 drug labels, providing the first scalable, interpretable benchmark for data-driven analysis of pediatric extrapolation.

To demonstrate the utility of the dataset, we built two baseline models, logistic regression and a fine-tuned BigBird transformer model [15], to benchmark extrapolation classification. Our goal is to provide a reproducible testbed that can support future work in pediatric drug development, clinical NLP, transfer learning and regulatory science.

Our main contributions are:

- **Regulator-grounded dataset**. PED-X-Bench converts previously unstructured FDA text into 737 machine-readable records, each linking an extrapolation label to its explicit rationale and supporting pediatric evidence.
- **Reasoning-model annotation pipeline**. We show that a reasoning-based LLM with 2-stage prompting can pre-label hundreds of FDA labels with expert-level fidelity, reducing manual effort by an order of magnitude.
- **Baseline benchmarks**. We benchmark this data with logistic regression and BERT-based BigBird transformer reference models to demonstrate its utility.

## 2 The *PED-X-Bench* Dataset

**PED-X-Bench** is the first openly available corpus that encodes FDA extrapolation decisions as a four-class classification task. The dataset comprises 737 labelling sections issued between 2007 and 2024 in which pediatric indications were considered. For each label we supply three artifact groups: (i) a categorical extrapolation label; Full, Partial, None or Unlabeled, derived from regulatory text; (ii) concise, large-language-model-generated summaries of pediatric efficacy, PK, and safety evidence that have been manually verified for accuracy as well as rationales for its decisions; and (iii) cleaned metadata that capture indication, approval year, brand and generic names, study design, sample size, age range, study centres, participating countries and, where BPCA trials are involved, race and ethnicity information.

**Category definitions** Our categorical definitions mirror regulatory practice. *Full* is assigned when disease progression, treatment response, and exposure–response are sufficiently concordant for pediatric PK (with or without targeted safety data) to substitute for efficacy evidence. *None* denotes clear dissimilarity that requires at least one adequate pediatric efficacy trial. *Partial* occupies the middle ground in which disease and therapeutic response appear similar but uncertainty persists about exposure–response concordance, prompting anything from a single pediatric trial to PK/PD bridging studies to confirm effect. *Unlabeled* applies when no pediatric data are available.

By transforming implicit FDA reasoning into explicit, machine-readable labels and anchoring each to its supporting text and metadata, PED-X-Bench provides a reproducible platform for tasks ranging from classification to evidence retrieval and policy analysis, and it lays a foundation for safer, evidence-based pediatric therapeutics.

## 3 Methods

### 3.1 Data source and preprocessing

To create PED-X-Bench, we began with the FDA spreadsheet that lists every pediatric labelling change triggered by BPCA, PREA, or the legacy pediatric Rule since 1998^1^. Each pediatric labeling change includes the date of the pediatric labeling change, specific drug or biological product, indication(s) studied, a summary of the labeling change, therapeutic category and type of legislation. It also contains pediatric study characteristics for the clinical trials conducted to support each pediatric labeling change, including the study number, type of study, study design, number of pediatric participants, ages studied, number of study centers, number of countries and, for BPCA clinical trials, any available racial and ethnic information.

From the most recent release we *(i)* removed veterinary or device entries, *(ii)* collapsed obvious reformulations, and *(iii)* retained the 737 human drugs whose English PDF labels were available. All PDFs were downloaded, converted to plain text with pdftotext, and lightly cleaned to preserve page breaks and tables.

### 3.2 LLM pipeline for extrapolating labels

To generate drug extrapolation categories from FDA labels, we devised a two-stage chain-of-thought pipeline using *o3-mini*. A *summary prompt* first scans the full label and extracts every sentence that cites pediatric efficacy, PK, or safety evidence. The resulting evidence block is passed to a *classification prompt* that *(i)* assigns one of four outcomes (Full, Partial, None, Unlabeled), *(ii)* provides the supporting rationale, and *(iii)* produces ≤150-word efficacy and safety summaries. Figure 2 illustrates both prompts.

**Figure 1:**
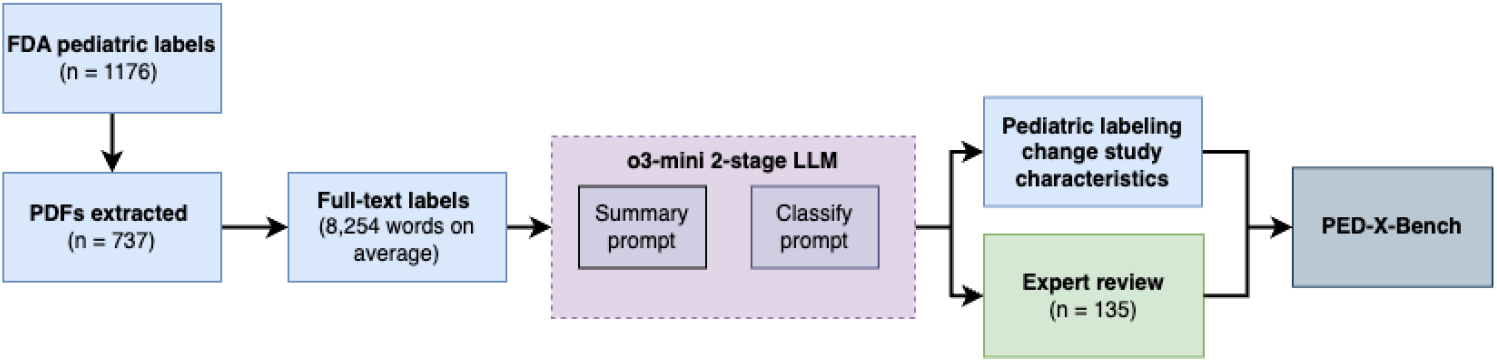
Two-stage LLM pipeline used to build *PED-X-Bench*. The model first extracts pediatric evidence from the full FDA label, then classifies the extrapolation outcome while generating concise efficacy and safety summaries. Expert review (green) provides the gold test set; study metadata (blue) are merged to form the final corpus.

**Figure 2:**
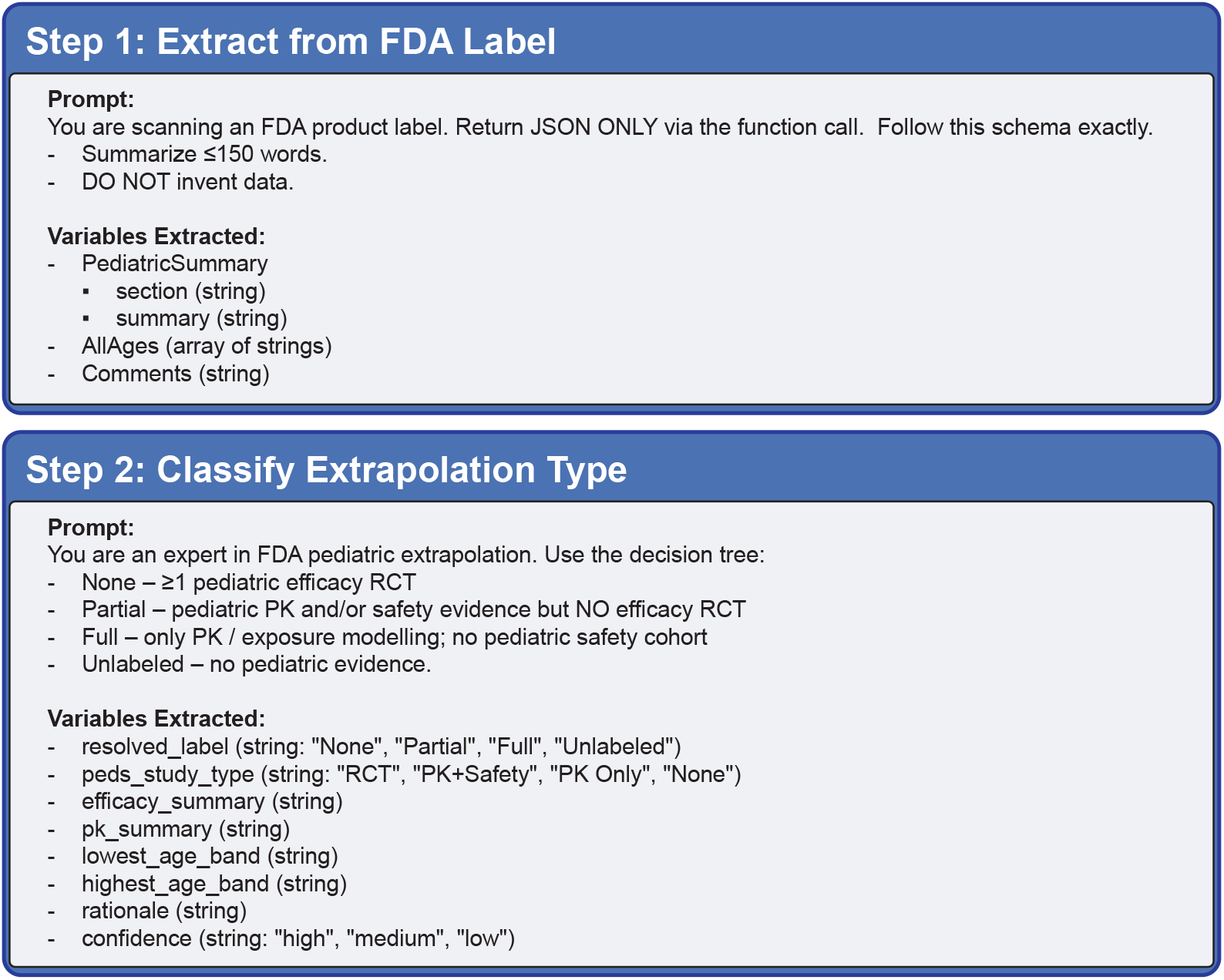
Summary and classification prompts used in the two-stage pipeline.

We performed three **ablation studies**: *(a)* a single-stage classifier over the full label; *(b)* the same two-stage pipeline run with *gpt-4o-mini*; and *(c)* the single-stage prompt plus a verifier that rereads the label and fixes JSON formatting errors.

### 3.3 Expert adjudication

For manual review, we recruited nine annotators, eight biomedical data scientists and one clinician, who independently reviewed 135 randomly sampled labels covering a broad therapeutic mix (antibacterials, antihistamines, anti-epileptics, asthma agents, oncology, antivirals). Annotators followed a written guide, recorded an extrapolation label, pasted verbatim efficacy- and safety-related evidence with page numbers, and flagged uncertainties. Inter-annotator agreement after consensus discussion reached κ = 0.72. These 135 labels form our gold-standard test-and-validation set; the remaining 602 machine-labelled records constitute silver-standard training data.

### 3.4 Baseline classifiers

We benchmark two complementary models on a fixed train / dev / test split of 687 / 85 / 34 labels.

#### Logistic regression on metadata

We z-scored the numeric features (patient counts, minimum and maximum age) and one-hot encoded the categorical ones (legislation type, therapeutic area), producing a 253-dimensional sparse feature vector for every label. A multinomial logistic regression with class-balanced weights was tuned on the development set for penalty (*𝓁*_1_, *𝓁*_2_, elastic-net), *C* ∈{0.01, 0.1, 1, 3, 10}, and *l*_1_ ratio, optimising macro-F1, then evaluated once on the test set.

#### BigBird on full text

We fine-tuned `google/bigbird-roberta-base` on tokenised label text truncated to 4096 tokens. raining used mixed precision, gradient checkpointing, and an effective batch size of eight (batch = 1, accumulation = 8) for four epochs with AdamW (lr = 2 × 10^*−*5^). The checkpoint with highest dev accuracy was re-evaluated on the test set.

Accuracy, macro-F1, cross-entropy loss, and ablation results appear in Section 4.

## 4 Results

Here we present the details about the PED-X-Bench dataset, then we discuss our validation with human annotated dataset and the effect of ablations on performance. Finally we illustrate the utility of our dataset as a benchmark in data extrapolation classification tasks.

### 4.1 Dataset overview

Figure 3A shows that *None* decisions dominate PED-X-Bench (60 %, 445/737), *Partial* accounts for 31 %, whereas *Full* borrowing is rare (3 %) and 6 % of records are *Unlabeled*. Outcome frequency co-varies with the legislative route (Fig. 3B): 409i (BPCA) submissions are overwhelmingly *None*, whereas PREA-only programmes obtain *Partial* or *Full* borrowing almost twice as often. Age coverage, rather than study size, distinguishes the classes (Fig. 3C–D): *Full* approvals span the entire pediatric spectrum, yet even they rarely cite more than three pediatric studies.

**Figure 3:**
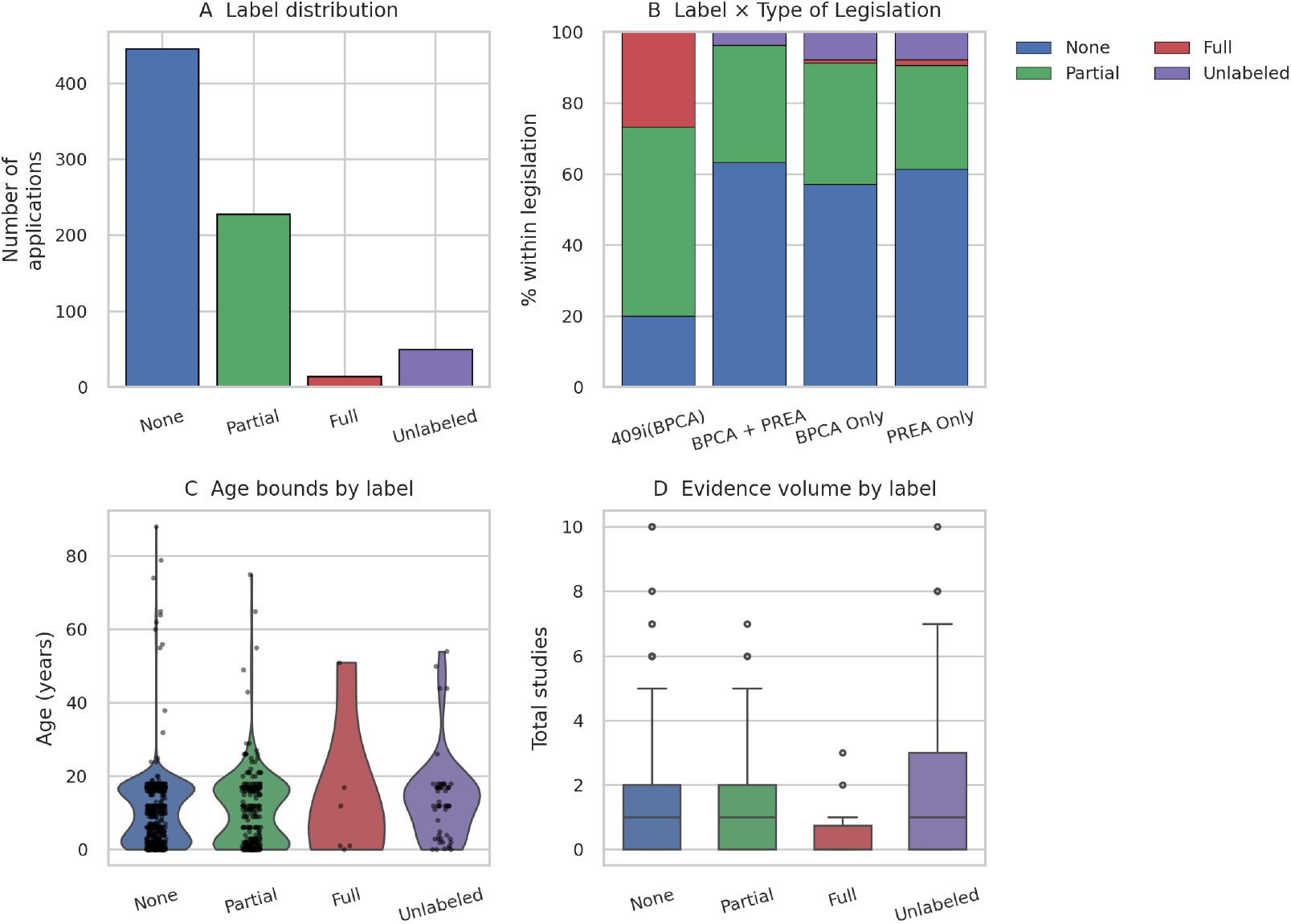
Corpus statistics. A) Label distribution. B) Extrapolation outcome by legislative pathway. C) Age-range coverage for each class (violins show min–max bounds). D) Number of pediatric studies cited per label.

Study-design flags (Fig. 4) reinforce this narrative. Classical efficacy and safety trials appear in *>*60 % of *None* labels but in *<*25 % of *Full*, indicating that when extrapolation is granted, sponsors rely primarily on pharmacokinetic/pharmacodynamic bridging rather than new randomised efficacy studies. Dose-escalation and neonatal studies remain scarce across the board.

**Figure 4:**
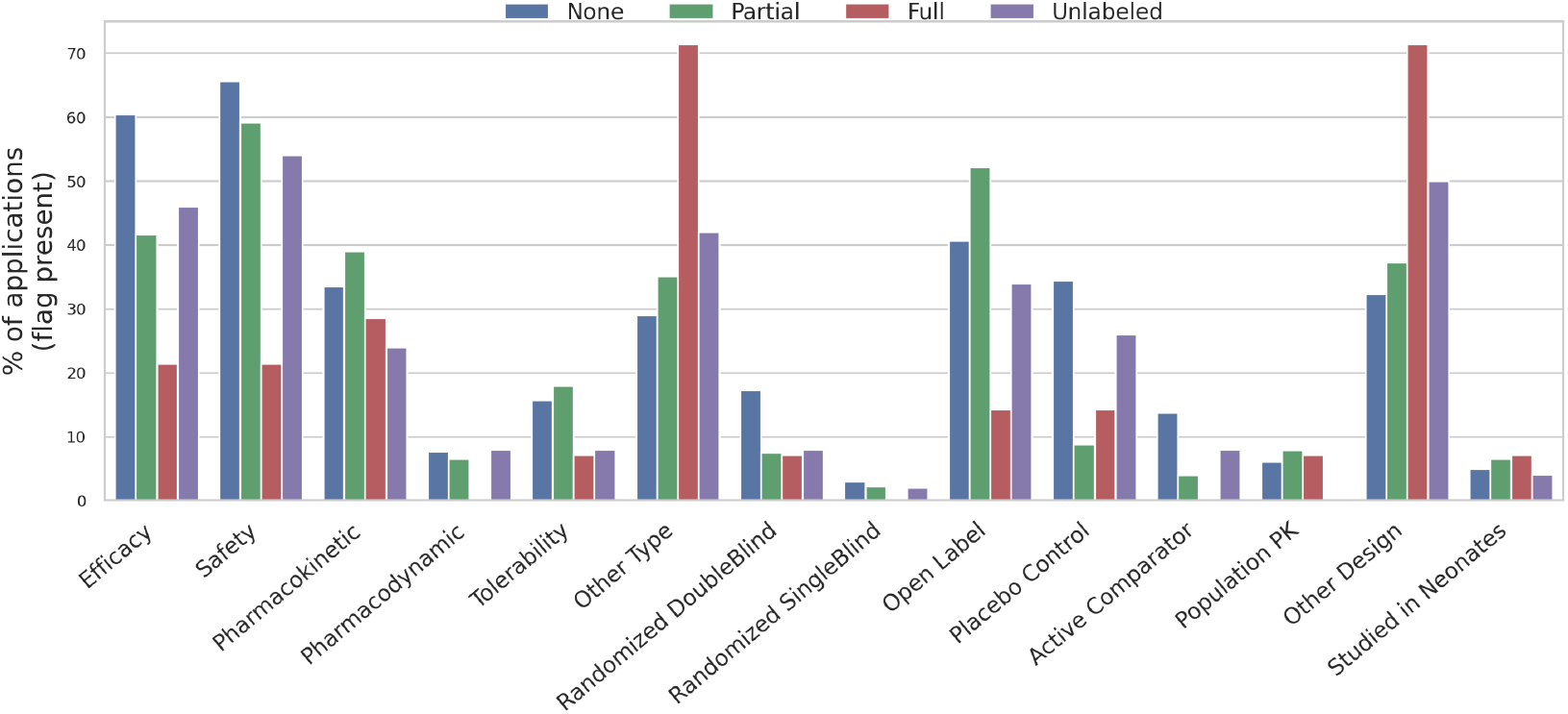
Study-design and outcome flags by extrapolation class. Bars denote the percentage of applications with each PLC flag.

### 4.2 Ablation studies on the LLM pipeline

Table 2 shows that swapping *o3-mini* for *gpt-4o-mini* in the two-stage pipeline drops accuracy from 0.74 to 0.53 and macro-F1 from 0.63 to 0.29, largely due to hallucinated rationales. A single-stage prompt recovers accuracy but lowers agreement with reviewers. Adding a verifier boosts accuracy to 0.80 but collapses many borderline cases into the majority class, lowering balanced F1. Hence we keep the two-stage *o3-mini* pipeline as the official labeller.

**Table 1:** Extrapolation categories used in *PED-X-Bench*.

| Category | Brief description |
| --- | --- |
| None | At least one adequate, well-controlled pediatric efficacy RCT conducted; no borrowing from adult data. |
| Partial | Adult efficacy RCT(s) accepted; pediatric PK ± safety studies bridge exposure–response or confirm dose. |
| Full | pediatric evidence limited to PK (or dosing equations) and safety; adult efficacy is fully borrowed. |
| Unlabeled | No pediatric data reported; extrapolation status not specified in the public label. |

**Table 2:** Ablation results for the LLM labelling pipeline.

| Variant | Accuracy | Macro-F1 | $\kappa$ |
| --- | --- | --- | --- |
| Two-stage (*o3-mini*) | 0.740 | **0.633** | 0.722 |
| Two-stage (*gpt-4o-mini*) | 0.534 | 0.294 | 0.602 |
| Single-stage (full label) | 0.725 | 0.637 | 0.619 |
| Single-stage + verifier | 0.804 | 0.538 | 0.706 |

### 4.3 Baseline classifiers

As shown in Table 3, a metadata-only logistic model matches BigBird’s accuracy while doubling its macro-F1, underscoring that coarse regulatory descriptors encode much of the decision boundary. Unlocking the full potential of long-sequence transformers will require larger, better-balanced corpora or objectives that respect the label hierarchy. BigBird processed ≈15 documents s^*−*1^ on a single A100 GPU, whereas the logistic model trained in under a second on CPU.

**Table 3:**
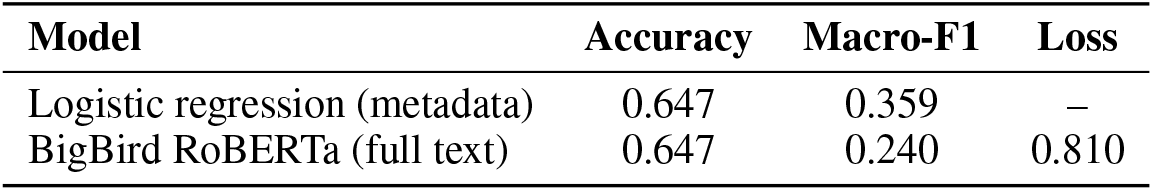
Baseline performance on the blind test split (34 labels).

| Model | Accuracy | Macro-F1 | Loss |
| --- | --- | --- | --- |
| Logistic regression (metadata) | 0.647 | 0.359 | – |
| BigBird RoBERTa (full text) | 0.647 | 0.240 | 0.810 |

## 5 Discussion

We presented *PED-X-Bench*, the first dataset that encodes FDA pediatric extrapolation outcomes as a four–way classification task directly from public labels. A two-stage chain-of-thought pipeline based on *o3-mini* labels the corpus at scale and reaches substantial agreement with nine expert reviewers (*κ* = 0.72).

PED-X-Bench opens a window onto two decades of FDA thinking about pediatric extrapolation. The aggregate statistics confirm long-suspected patterns—complete borrowing is exceptional, partial borrowing is closely tied to PREA submissions, and neonatal evidence remains sparse. But they also quantify the magnitude of those skews with far greater resolution than the handful of earlier manual audits. By coupling every categorical outcome to its verbatim rationale and to harmonized study metadata, the corpus turns a once-opaque regulatory process into a set of tractable machine-learning tasks. The initial experiments show that lightweight logistic regression already rivals a transformer fed the full label text, implying that much of the decision boundary is summarized by a few high-level descriptors. In other words, today’s models have not yet learned to exploit the narrative subtleties that regulators weigh when they decide how far adult evidence can stretch.

That observation cuts both ways. On the one hand, it underscores the value of simple baselines and cautions against assuming that ever-larger language models automatically outperform structured features. On the other hand, it signals clear headroom: with balanced data, hierarchy-aware objectives, or retrieval-augmented inputs that surface analogous historical decisions, long-sequence transformers should be able to integrate fine-grained clinical nuance and outperform the metadata-only baseline. PED-X-Bench also invites multimodal approaches that blend external evidence such as clinical-trial registries, population-PK simulations, real-world safety reports, and disease-similarity embeddings so that models can reason over data unavailable in the label itself. The corpus therefore functions both as a benchmark and as an incubator for new methods in hierarchical text classification, evidence retrieval from multi-modal sources, and explanation generation.

Several limitations deserve emphasis. First, the dataset inherits the biases of publicly available labels: negative trials, interim analyses, and unpublished regulatory correspondence are absent, so PED-X-Bench necessarily reflects the final, public-facing narrative rather than the full deliberative record. Second, class imbalance is severe, with only 22 fully extrapolated cases; while this mirrors real-world rarity it also constrains model learning. Third, extrapolation decisions evolve as new pharmacology or real-world safety data emerge; our annotations capture a single snapshot (2007-2024) and will require periodic refreshes to stay current. And finally, the current annotations address extrapolation at the level of the entire product rather than at the granularity of individual indications or dosage forms, a simplification that future work could refine.

Looking forward, we view PED-X-Bench as the foundational layer of a broader research agenda. The next step is to build multi-agent systems that draw simultaneously on clinical-trial registries, population-PK simulations, disease-similarity graphs, and real-time literature feeds to generate end-to-end extrapolation recommendations. The corpus provides both the supervision signals and the evaluation scaffolding for such agents: any proposed recommendation must map back onto the four regulatory outcomes and cite evidence spans comparable to those captured here. Beyond modeling, the dataset can help regulators audit consistency across therapeutic areas, identify edge cases, such as neonates, where evidence remains thin, and prioritize future incentive programs.

Finally, there is a broader impact dimension. If successful, automated extrapolation tools could accelerate pediatric drug development by highlighting where adult data already suffice and where targeted trials are indispensable, thereby reducing ethical burdens and costs. At the same time, algorithmic recommendations must remain transparent, auditable, and subject to expert oversight; PED-X-Bench’s linkage of predictions to textual rationales is a step in that direction. We hope this resource catalyzes a dialogue between the machine-learning and regulatory communities, ultimately moving pediatric therapeutics toward faster, safer, and more evidence-grounded approvals.

## 6 Conclusion

PED-X-Bench transforms two decades of FDA labelling into the first open, machine-readable resource for studying pediatric extrapolation, coupling four-way outcome labels to their precise textual rationales and rich study metadata. Benchmark experiments show that simple metadata already explains much of the regulatory boundary, while leaving clear headroom for more innovative methods. By providing a rigorously curated corpus, baseline scores, and an extensible LLM annotation pipeline, we lay the groundwork for future systems that can reason across trials, pharmacology, and real-world evidence to deliver transparent, data-driven extrapolation recommendations and thereby, advancing both clinical NLP research and pediatric drug development.

## Data Availability

All data produced are available online at https://huggingface.co/datasets/apoorvasrinivasan/Ped-X-Bench

## Footnotes

1 https://www.fda.gov/science-research/pediatrics/pediatric-labeling-changes

